# Quality of life and health-related utility after head&neck cancer surgery

**DOI:** 10.1101/2021.01.08.20249019

**Authors:** Enea Parimbelli, Christian Simon, Federico Soldati, Lorry Duchoud, Gian Luca Armas, John de Almeida, Silvana Quaglini

## Abstract

**Purpose:** This work describes the methodology adopted and the results obtained in a utility elicitation task. The purpose was to elicit utility coefficients (UCs) needed to calculate quality-adjusted life years for a cost/utility analysis of TORS (Trans-Oral robotic Surgery) versus TLM (Trans-oral Laser Microsurgery), which are two minimally-invasive trans-oral surgery techniques for head & neck cancers.

**Methods:** Since the economic evaluation would be conducted from the point of view of the Swiss healthcare system, Swiss people (healthy volunteers) have been interviewed in order to tailor the model to that specific country. The utility elicitation was performed using a computerized tool (UceWeb). Standard gamble and rating scale methods were used.

**Results:** UCs have been elicited from 47 individuals, each one providing values for 18 health states, for a total of 1692 expected values. Health states, described using graphical factsheets, ranged from remission to palliative care. Elicited UCs were different among states, ranging from 0.980 to 0.213. Those values were comparable to previously published results from a Canadian population, except for states related to recurrent disease (local, regional, and distant), and palliation, where the Swiss population showed lower utility values.

**Conclusion:** From a methodological point of view, our study shows that the UceWeb tool can be profitably used for utility elicitation from healthy volunteers. From an application point of view, the study provides utility values that can be used not only for a specific cost-utility analysis, but for future studies involving health states following trans-oral head & neck surgery. Moreover, the study confirms that some UCs vary among countries, demanding for tailored elicitation tasks.

## 1. Introduction

One of the major goals in surgery is to preserve function. In head and neck surgery open approaches have long been favored, but more recently minimally-invasive techniques have been introduced to reduce access-related morbidity and consecutive functional deficits. Such novel approaches are mostly used to access cancers in the pharynx in particular with trans-oral approaches. Nowadays two techniques are mainly used, i.e. trans-oral laser microsurgery (TLM) and trans-oral robotic surgery (TORS). While the philosophy of both techniques is similar, the costs related to equipment and surgical strategy are different, demanding an economic and health state related evaluation to better judge which technique to pursue.

In the past few years, we have developed a model for performing such an economic evaluation [1]. The model has been developed as a decision tree [2], using the TreeAge Pro software [3]. It represents the options to be compared (TORS and TLM) and their possible consequences in terms of probability of occurrence (complications of the intervention, additional treatments needed, relapse, remission, etc.). Patients’ transitions among different health states are represented through Markov processes [4]. Direct costs associated with every intervention and every health state treatment have also been defined.

From such a model, expected values of survival and costs for each treatment option may be estimated. However, among the possible types of economic evaluations for healthcare programs, we were interested in Cost/Utility analysis (CUA), because it also takes into account the patients’ quality of life.

CUA, which will be better described in the next section, requires weighting survival with so-called utility-coefficients (UCs), i.e. a measure of the desirability of health states. This measure is highly subjective, and may vary among different populations, depending (also) on cultural, geographical, and economical aspects [5,6]. Thus, in principle, to make the CUA consistent, UCs should be elicited from the same population targeted by the specific economic evaluation study, as well as it is a common practice to use a specific country’s costs. Unfortunately, utility elicitation is a very time consuming and challenging task, and often researchers rely on values found in the literature, even if these values have been collected from different populations. To overcome this methodological pitfall, since our overarching goal was to perform a CUA from the point of view of the Swiss health care system, the model quantification would take into account the preferences of the Swiss population.

The aim of this paper is first of all to present the methodology adopted for eliciting those preferences. Secondly, results will be shown about the UCs that will value the health states represented in the decision tree, i.e. the health states patients typically experience after head & neck cancer surgery.

In the following, we present in a deeper detail the CUA methodology, then we discuss the existing literature about UCs in head & neck cancer, we describe the tool used for utility elicitation, the population involved, the aids used for the interviews and the results.

### 1.1 Related work

A very comprehensive, systematic literature review of health state utility values in head and neck cancer was published in 2017 [7]. However, only one study, namely de Almeida et al [8] reported UCs related to a number of health states following TORS or radiotherapy in oropharyngeal cancer patients. Also, none was referring to a Swiss population. The review concludes that “…There is currently a lack of research for some disease phases including recurrent and metastatic cancer, and treatment-related complications”.

After 2017, additional papers have been published on the argument. Some studies, such as in [9], investigated face and construct validity of direct utility elicitation methods. Liao et al [10], comparing utility values in Taiwan and Sweden, confirm the need of understanding population differences and the same claim is reported in [11]: “Future health state valuations would ideally be based on representative value sets to reflect the different populations within and beyond the currently evaluated countries.” Again, no study targeted Swiss population, and no study reported UCs for the health states after mini-invasive surgery.

Our work contributes to fill the gap by eliciting UCs directly from a Swiss population using direct methods, and including tumor recurrences, metastatic phase and treatment complications.

## 2. Methods

### 2.1 Cost/Utility analysis and utility coefficients

Cost/Utility analysis (CUA) is a kind of economic evaluation of healthcare programs, comparing two or more options in terms of costs and benefits. In CUA, Quality-Adjusted Life Years (QALYs) are commonly used as the primary health outcome [12]. Sometimes, one of the options *dominates* the other ones, i.e., it is more effective in terms of QALYs and less expensive. On the contrary, when it turns out that the most effective option is also the most expensive, a popular indicator is ICUR (Incremental Cost/Utility Ratio), representing the cost per QALY gained. ICUR may be compared to existing agreed-on thresholds to judge whether or not the option is worth to be implemented [13,14].

To calculate QALYs, a patient’s expected survival is split in time intervals, each one (Ti) spent in a specific condition Ci. Assuming that Ci is associated with a utility coefficient UCi, QALYs are computed as the weighted sum of Ti x UCi. The UCs are numbers ranging from 0 to 1, where 0 is for the worst possible health state (usually death) and 1 is for the best possible one (perfect health). An UC conveys the patient’s feeling about the goodness of a health state.

Different methods have been proposed in the literature for eliciting UCs. The standard gamble (SG) is the soundest one from the theoretical point of view, since it derives directly from the axioms of the utility theory [15]. In SG, to elicit the UC for a sub-optimal state S, the interviewee must enter a hypothetical scenario, and choose between living his expected life in the S state or accepting a gamble whose outcomes are complete healing or sudden death with probability p. The gamble is usually presented as a hypothetical treatment (e.g., a surgical intervention or a drug) able to cure the patient, but presenting a risk *p* of death, due for example to intraoperative complications or drug toxicity. The first risk value proposed can be arbitrary (e.g., 0.5), and then varied (increased or decreased according to the answers, so-called *ping-pong* procedure) until the interviewee can’t decide about the bet. At that point, the UC is computed as (1–p). The rationale is that if S is a very bad state, the individual should accept a treatment even when the treatment-related risk is high. Being the UC calculated as 1-p, a bad state turns out to have a low UC. On the other hand, if a health state is considered similar to the perfect health, no risk will be accepted (or a risk value=0), and then the UC will be 1.

While being theoretically sound, SG is not always easy to administer, because it requires the interviewee to enter the *hypothetical scenario*, to have a clear notion of *risk*, and to understand the *gamble meaning*.

Among other methods, the simplest one is Rating Scale (RS). A scale between 0 and 100 is usually presented to the interviewee, asking him to position the S state within the scale. As a drawback to its simplicity, the value obtained is not a true UC, but it is useful to test the user confidence with the interview and as a starting point for further elicitation with sounder methods. For example, it can be used to assess the initial risk value shown in the SG.

A complete review of the utility elicitation methods is beyond the scope of this paper and can be found in [16].

In our study we used both SG and RS for all the utility elicitations. As described in sections 4 and 5, elicitations were supported by a computerized tool and factsheets describing in detail the health states for which UCs should be elicited.

### 2.2 Study population

Whether patients or healthy individuals of the general population should provide UCs to be incorporated into a CUA has been debated in the literature. Despite patients, of course, are more aware about their health state and how it affects their life, the general population is probably more suitable for analyses that will be used by healthcare policy makers to take decisions that will affect the population as a whole. One motivation for relying in general community values instead of patients’ ones is that patients may develop adapting behaviors to live with a certain heath state, thus overestimating the true state utility [11].

As a matter of fact, there is a number of recommendations to use informed general population values in decision making [16–18], among which the United States Public Health Services panel’s [19].

On this line, we interviewed a set of 71 healthy volunteers from Switzerland. While not being patients, they are in a daily contact with patients, and thus they are in principle more informed than the general population. However, some of them may have less familiarity with patients and specific conditions. Thus, we decided to prepare a series of factsheets describing all the health states considered in the study (see the Methods section and additional material). The preparation of the scenario sheets involved feedback-refinement iterations among the clinical experts from CHUV. The final version of the sheets has been used during the interviews with all the volunteers.

To calculate the interviewees’ sample size, we considered estimating the mean value of utility coefficients with a precision (margin of error) of 0.05, assuming a standard deviation of 0.15. To achieve a confidence level of 95%, the minimum number of persons needed is 38. In other words, if we select a random sample of 38 individuals from a population, and determine the mean to be say y, we would be 95% confident that the mean in the population lies between (y - 0.05) and (y + 0.05) [20]. To be conservative, and to prevent loss of statistical power due to drop-out or data issues, we enrolled 47 volunteers.

### 2.3 The computerized tool for utility elicitation

Utility elicitation is a challenging task. In order to obtain consistent values from all the individuals, it is important that everybody is interviewed following the same procedure. Using a computer-based tool helps the interviewers to pursue this objective. We used UceWeb, a system that we developed in the last years [1,21,22] and that has been already validated in other medical contexts [23,24]. The distinguishing features of UceWeb are:

- a graphical interface implementing several methods for utility elicitation;
- a decision support facility suggesting the best elicitation method according to the interviewee’s and health state characteristics;
- a common terminology (SNOMED) for the health states for which UCs are elicited (additional states, not covered by SNOMED, may be added if necessary);
- a collaborative environment where different researchers can feed the UCs repository, while providing a basic profile for every interviewed individual (age, gender, country, if he is a patient or not, and other features known to affect an individual’s preferences). This will allow future studies to have larger and larger sets of UCs for specific target populations. The study described in this paper contributed to this repository indeed;
- a direct link to TreeAge Pro, allowing to run a decision tree just after having elicited UCs from a single patient or having retrieved a UC set for a target population from the repository.

UceWeb is a multiuser (and multilingual) system and this allowed the interviewers to work in parallel and save time. For the current study, French language has been used.

### 2.4. Utility elicitation sessions

The elicitations were done in 9 sessions over a 101 days period.

As anticipated in the previous section, all the interviews were done using UceWeb and supported by the same set of factsheets describing the clinical context, the treatments and their possible consequences. In this way, all the volunteers received the same information in the same format. Two physicians (FS and LD) performed all the interviews and answered all the volunteers’ questions. First of all, the clinical problem was described, relying on [25],[26,27], as in the following (we report the same words used for informing the volunteers, which is useful for the reader as well, to understand the context, while omitting some details for sake of space).

### 2.5 The clinical problem (as explained at the beginning of each interview)

Oropharyngeal carcinomas (OPCs) represent a major health problem. The oropharynx is the region of the throat behind the mouth. It contains, as main structures, the two palatine tonsils on each side and the base of the tongue. These areas play a major role in swallowing and breathing. Unfortunately, this region can be the site of cancer. The most common cancer found in the oropharynx is squamous cell carcinoma. … Patients with OPC associated with HPV tend to be young, non-smoking men.

The possible treatments

The type of treatment that can be offered to patients suffering from OPC is of three types: a surgical intervention which removes the cancerous tissue, radiotherapy (RT), which induces the death of cancer cells, and finally chemotherapy (CT) in the form of a medicine (taken intravenously) that attacks cancer cells all over the body through the bloodstream. How these treatment modalities are chosen and combined depends on the stage of the cancer, the availability of treatment, and the patient’s preferences. The most common methods and combinations are:

- Surgery only
- Radiotherapy only
- Radiotherapy and Chemotherapy (Primary concurrent chemoradiation - CCRT)
- Surgery followed by radiotherapy
- Surgery followed by CCRT

When radiotherapy or chemo-radiation are added after surgery, this treatment is called “adjuvant”. In the case of adjuvant radiation therapy, the radiation dose is reduced compared to primary radiation therapy alone.

In order to improve functional recovery after surgery, i.e. your ability to eat, breathe and speak normally, new techniques have been developed. They consist of fully endoscopic approaches to the tumour by mouth, thus avoiding access through the neck and reducing the morbidity associated with access. This leads to a much faster recovery after surgery and a better functional result. The tumour is visualized with endoscopes and microscopes.

The two main endoscopic techniques practiced today are trans-oral robotic surgery (TORS) and trans-oral laser microsurgery (TLM). For TORS, a retractor is used to open the mouth to create space for the robotic camera and surgical instruments. Then, the surgeon uses a surgical robot to view and access the structures of the pharynx (back of the throat). The tumour is removed with an electric knife. In TLM, retractors are also used to open the mouth and access the throat, but with this technique, the surgeon uses a microscope and a laser to remove the tissue.

Both techniques show pros and cons. When TLM is performed, the visual field is usually quite small …this can compromise the correct assessment of surgical margins and trigger unnecessary adjuvant therapy. However, the precision of TLM is exceptionally high ….

TORS, on the contrary, allows for resection in one piece based on better visualization with the available endoscopes and cutting instruments…. Therefore, the analysis of surgical margins is more precise. On the contrary, the accuracy of the dissection, particularly at the deep margins, may not be as satisfactory as with the laser and the microscope.

Surgical approaches to the oropharynx are generally associated with dissection of the neck lymph nodes in order to remove the nodes eventually carrying tumour cells. An incision in the neck is made to allow the surgeon to remove the lymph nodes potentially contaminated with cancer cells.

### 2.6 Introducing the interview

After describing the clinical problem, the interview is introduced through a hypothetical *shared decision-making framework:*

“Imagine being a patient who consults an otolaryngologist following the diagnosis of cancer of the oropharynx. Your doctor will describe different treatment options and you will finally choose the option that best suits your wishes. After sufficient discussion with him, you will have enough information about the risks and benefits to be able to make a decision.”

After this introduction, the elicitation exercise begins. First of all, the oncologist describes the scenario #1, depicting the first month consequences of the surgical intervention, which are the same for TORS and TLM, and then he describes all the complications and further treatments that are considered in the decision model, for a total of 18 health states, which are reported in table 2:

**Table 1.**
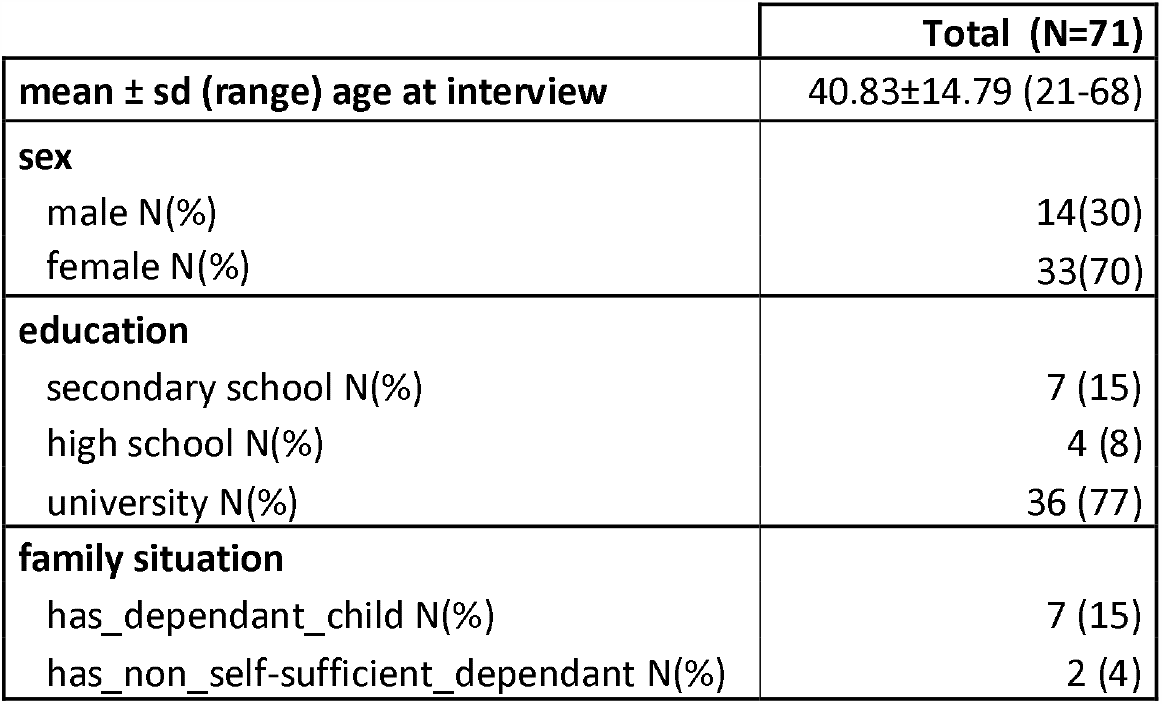
summarizes the interviewees’ characteristics.

|  | Total (N=71) |
| --- | --- |
| mean ± sd (range) age at interview | 40.83±14.79 (21-68) |
| **sex** |  |
| male N(%) | 14(30) |
| female N(%) | 33(70) |
| **education** |  |
| secondary school N(%) | 7 (15) |
| high school N(%) | 4 (8) |
| university N(%) | 36 (77) |
| **family situation** |  |
| has_dependant_child N(%) | 7 (15) |
| has_non_self-sufficient_dependant N(%) | 2 (4) |

**Table 2.** The health states to be valued with UCs

| Health state ID | State label | SNOMED code(s)* |
| --- | --- | --- |
| 1 | TLM) / TORS and lymph node resection | 10724008\|Microsurgery\|118438002\|Transoral approach\| |
| 2 | The same as above, but for a re-intervention | 64695001\|Repeat elective\|10724008\|Microsurgery\|118438002\|Transoral approach\| |
| 3 | Radiotherapy (RT) after surgery | 169351001\|Radiotherapy: infuse head/neck\|262061000\|Postoperative\| |
| 4 | Chemo-radiotherapy (CRT) after surgery | 169400008\|Chemo-radiotherapy: |
|  |  | IV\|262061000\|Postoperative\| |
| 5 | Tracheotomy long duration | 48387007\|Tracheotomy\| |
| 6 | Gastrostomy | 54956002\|Gastrostomy\| |
| 7 | Pharyngo-cutaneous fistula | 232413009\|Pharyngocutaneous fistula\| |
| 8 | Hospital re-admission for febrile neutropenia | 409089005\|Febrile neutropenia\| |
| 9 | Esopharyngeal stenosis | 232372008\|Nasopharyngeal stenosis\| |
| 10 | Osteo-radio-necrosis | 109333005\|Osteoradionecrosis\| |
| 11 | Post-operative hemorrhages | 110265006\|Postoperative hemorrhage\| |
| 12 | Remission after TORS/TLM | 277022003\|Remission\|10724008\|Microsurgery\|118438002\|Transoral approach\| |
| 13 | Remission after TORS/TLM + adjuvant | 277022003\|Remission\|10724008\|Microsurgery\|118438002\|Transoral approach\|169400008\|Chemo-radiotherapy: IV\| |
| 14 | Local recurrence (intervention needed) | 25173007\|Recurrent tumor\|255470001\|Local\|64695001\|Repeat elective\| |
| 15 | Local recurrence (RT or CRT needed) | 25173007\|Recurrent tumor\|255470001\|Local\|169400008\|Chemo-radiotherapy: IV\| |
| 16 | Regional recurrence | 25173007\|Recurrent tumor\|410674003\|Regional\| |
| 17 | Distant recurrence | 25173007\|Recurrent tumor\|261007001\|Distant\| |
| 18 | 18- Palliative care | 103735009\|Palliative care\| |

\*post-coordination[28] has been used for states that are not natively covered in SNOMED

As an example, the following box reports the factsheet for scenario #4 (the other ones are available in the additional material, all of them have been proposed to the volunteers in the French version):

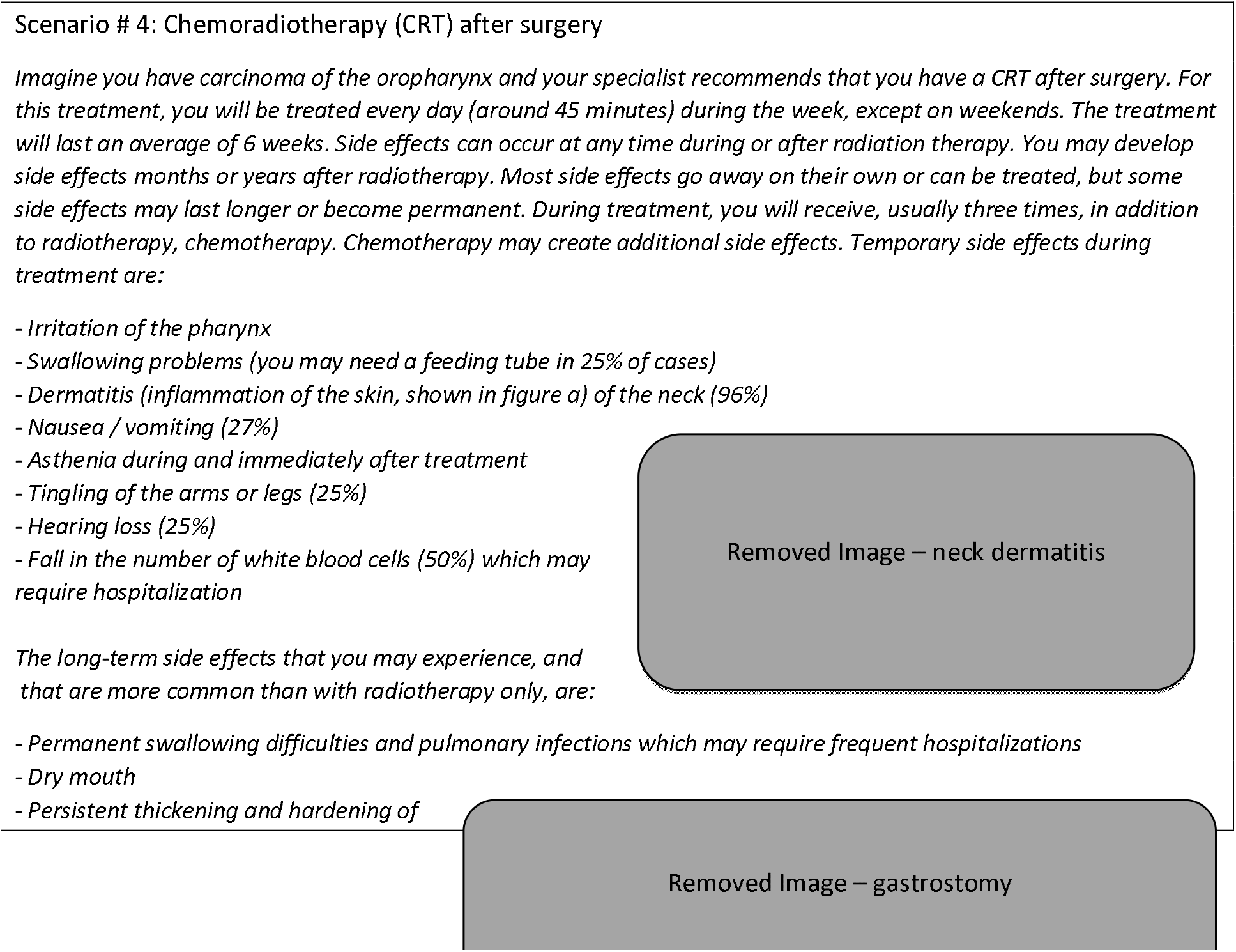
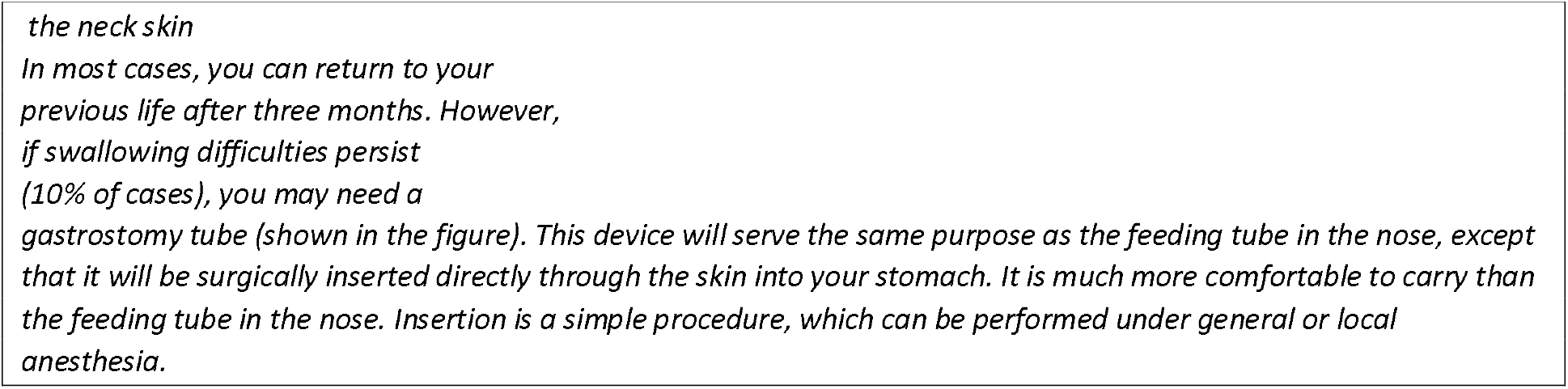

After explaining each scenario and addressing possible doubts of the interviewee, the following questions are asked, implementing the rating scale and standard gamble methods, respectively:

1. *On a scale of 0 to 100, where 0 represents death and 100 represents perfect health, where would you place the above scenario?* (Let UCrs be the UC elicited with this first question)
2. *Now imagine that you can choose between an adjuvant CRT and a pill that, taken at home, has the same effectiveness as the adjuvant CRT, but without side effects. However, taking the pill carries a risk of sudden death (we will go shortly in the value of the risk). Would you consider taking the pill if the risk is low enough?*

According to the standard gamble procedure, if the person does not accept even a very small risk, the interview is stopped, and the UC of the described state turns out to 1. On the contrary, if the person accepts the gamble, the first risk value proposed is (1-UCrs). As described in [21], this initialization shortens the SG ping-pong procedure.

The above two questions are the same for all the 18 scenarios, resulting in the RS and SG values for each health state.

## 3 Results

As mentioned, 47 Swiss individuals were enrolled for the study. A complete interview took from 18 to 69 minutes (43.5 ±12.5 minutes). Table 3 shows the summary statistics of the SG-elicited UCs and RS values for the different scenarios. We report mean and standard deviation to facilitate comparison with other possible values reported in the literature, and median and quartiles to reflect the non-normal distribution very skewed towards 1 for the SG method.

**Table 3.** The values elicited with standard gamble and rating scale methods

| ID | Scenario | Standard Gamble (UCs) |  |  |  |  |  |  | Rating Scale |  |  |  |  |  |  |
| --- | --- | --- | --- | --- | --- | --- | --- | --- | --- | --- | --- | --- | --- | --- | --- |
|  |  | mean | sd | max | min | median | q1 | q3 | mean | sd | max | min | median | q1 | q3 |
| 1 | TLM/TORS intervention (first month) | 0.902 | 0.203 | 1.000 | 0.245 | 1.000 | 0.968 | 1.000 | 0.546 | 0.200 | 1.000 | 0.100 | 0.510 | 0.395 | 0.725 |
| 2 | Re-intervention | 0.872 | 0.249 | 1.000 | 0.075 | 1.000 | 0.919 | 1.000 | 0.441 | 0.192 | 0.790 | 0.070 | 0.400 | 0.290 | 0.568 |
| 3 | RT after intervention | 0.850 | 0.275 | 1.000 | 0.040 | 1.000 | 0.880 | 1.000 | 0.448 | 0.193 | 0.840 | 0.050 | 0.400 | 0.300 | 0.630 |
| 4 | CRT after intervention | 0.794 | 0.317 | 1.000 | 0.030 | 1.000 | 0.548 | 1.000 | 0.334 | 0.172 | 0.650 | 0.030 | 0.300 | 0.200 | 0.500 |
| 5 | Tracheotomy long duration | 0.852 | 0.271 | 1.000 | 0.020 | 1.000 | 0.895 | 1.000 | 0.406 | 0.189 | 0.800 | 0.080 | 0.400 | 0.255 | 0.500 |
| 6 | Gastrostomy | 0.916 | 0.209 | 1.000 | 0.095 | 1.000 | 0.990 | 1.000 | 0.501 | 0.214 | 0.900 | 0.070 | 0.550 | 0.350 | 0.665 |
| 7 | Pharyngo-cutaneous fistula | 0.932 | 0.194 | 1.000 | 0.025 | 1.000 | 1.000 | 1.000 | 0.496 | 0.205 | 0.850 | 0.100 | 0.500 | 0.345 | 0.680 |
| 8 | Hospital re-admission for febrile neutropenia | 0.954 | 0.140 | 1.000 | 0.330 | 1.000 | 1.000 | 1.000 | 0.449 | 0.211 | 0.840 | 0.080 | 0.460 | 0.275 | 0.615 |
| 9 | Esopharyngeal stenosis | 0.826 | 0.284 | 1.000 | 0.000 | 0.975 | 0.833 | 1.000 | 0.322 | 0.180 | 0.790 | 0.000 | 0.300 | 0.185 | 0.420 |
| 10 | Osteo-radio-necrosis | 0.791 | 0.302 | 1.000 | 0.005 | 0.915 | 0.758 | 1.000 | 0.274 | 0.164 | 0.620 | 0.050 | 0.250 | 0.125 | 0.400 |
| 11 | post-operative hemorrhages | 0.910 | 0.203 | 1.000 | 0.120 | 1.000 | 0.970 | 1.000 | 0.542 | 0.204 | 0.950 | 0.120 | 0.500 | 0.395 | 0.680 |
| 12 | remission after TORS/TLM | 0.980 | 0.099 | 1.000 | 0.340 | 1.000 | 1.000 | 1.000 | 0.763 | 0.177 | 1.000 | 0.290 | 0.800 | 0.695 | 0.900 |
| 13 | Remission after TORS/TLM + adjuvant | 0.957 | 0.151 | 1.000 | 0.125 | 1.000 | 1.000 | 1.000 | 0.620 | 0.174 | 0.920 | 0.200 | 0.650 | 0.500 | 0.700 |
| 14 | Local recurrence (intervention needed) | 0.755 | 0.316 | 1.000 | 0.005 | 0.900 | 0.525 | 1.000 | 0.250 | 0.149 | 0.650 | 0.000 | 0.240 | 0.150 | 0.350 |
| 15 | Local recurrence (RT or CRT needed) | 0.771 | 0.302 | 1.000 | 0.005 | 0.935 | 0.598 | 1.000 | 0.264 | 0.134 | 0.620 | 0.000 | 0.250 | 0.160 | 0.355 |
| 16 | Regional recurrence | 0.859 | 0.283 | 1.000 | 0.005 | 1.000 | 0.878 | 1.000 | 0.378 | 0.188 | 0.790 | 0.030 | 0.350 | 0.250 | 0.500 |
| 17 | Distant recurrence | 0.307 | 0.350 | 1.000 | 0.000 | 0.100 | 0.015 | 0.513 | 0.108 | 0.109 | 0.500 | 0.000 | 0.100 | 0.040 | 0.130 |
| 18 | Palliative care | 0.213 | 0.336 | 1.000 | 0.000 | 0.010 | 0.005 | 0.320 | 0.072 | 0.086 | 0.400 | 0.000 | 0.050 | 0.015 | 0.090 |

## 4. Discussion

### 4.1 Ranking of health states

Considering the mean values, the 18 health states were ranked in the same order by the two methods 9 out of 18 times. Table 4 reports the ordering of elicited UCs from lowest (e.g. palliative care) to highest (remission). As shown, the two methods rank in the same order the 5 worst and the 2 best states, so showing higher agreement for poorest states. This is reasonable, given the already mentioned skeweness of SG values towards high values.

**Table 4.**
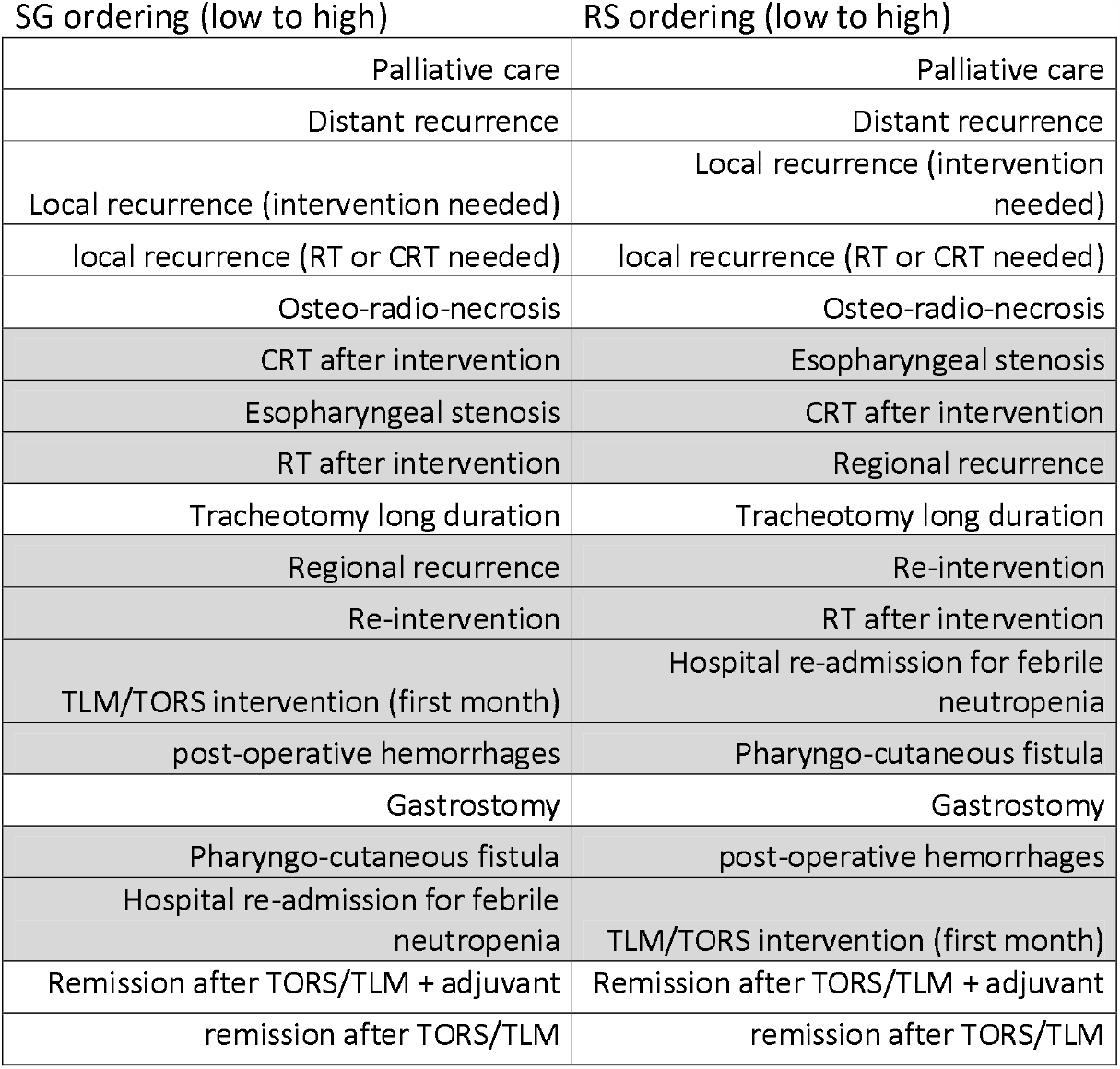
Ranking of the different health states according to values elicited with SG and RS. Grey background highlights the differences in ranking.

### 4.2 Correlation of elicited values with interviewees’ profile

Considering all the elicited values, RS and SG values showed a good correlation (Figure 1).

**Figure 1.**
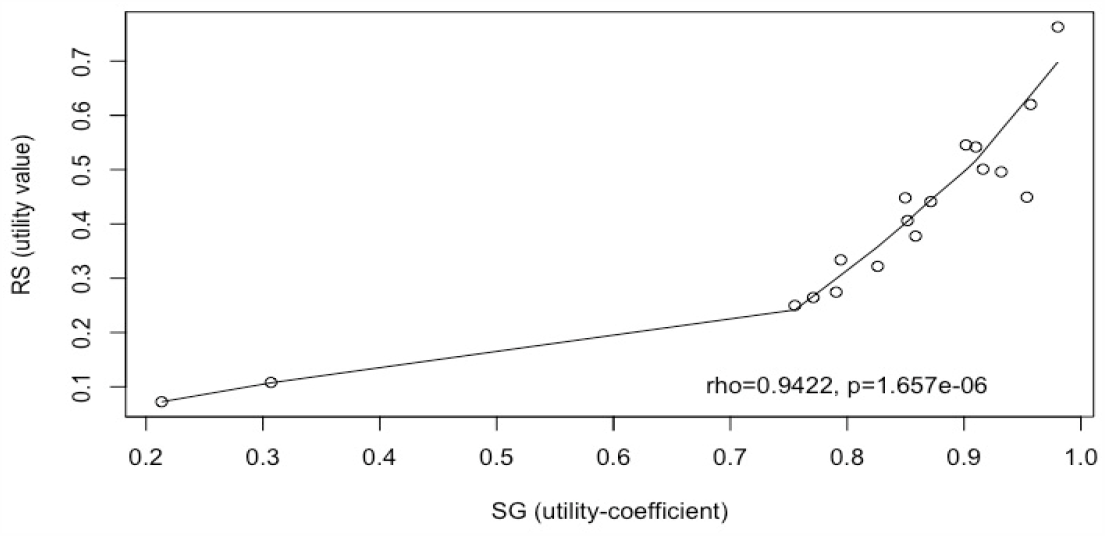
Correlation plot of mean SG utilities and RS values.

**Figure 2.**
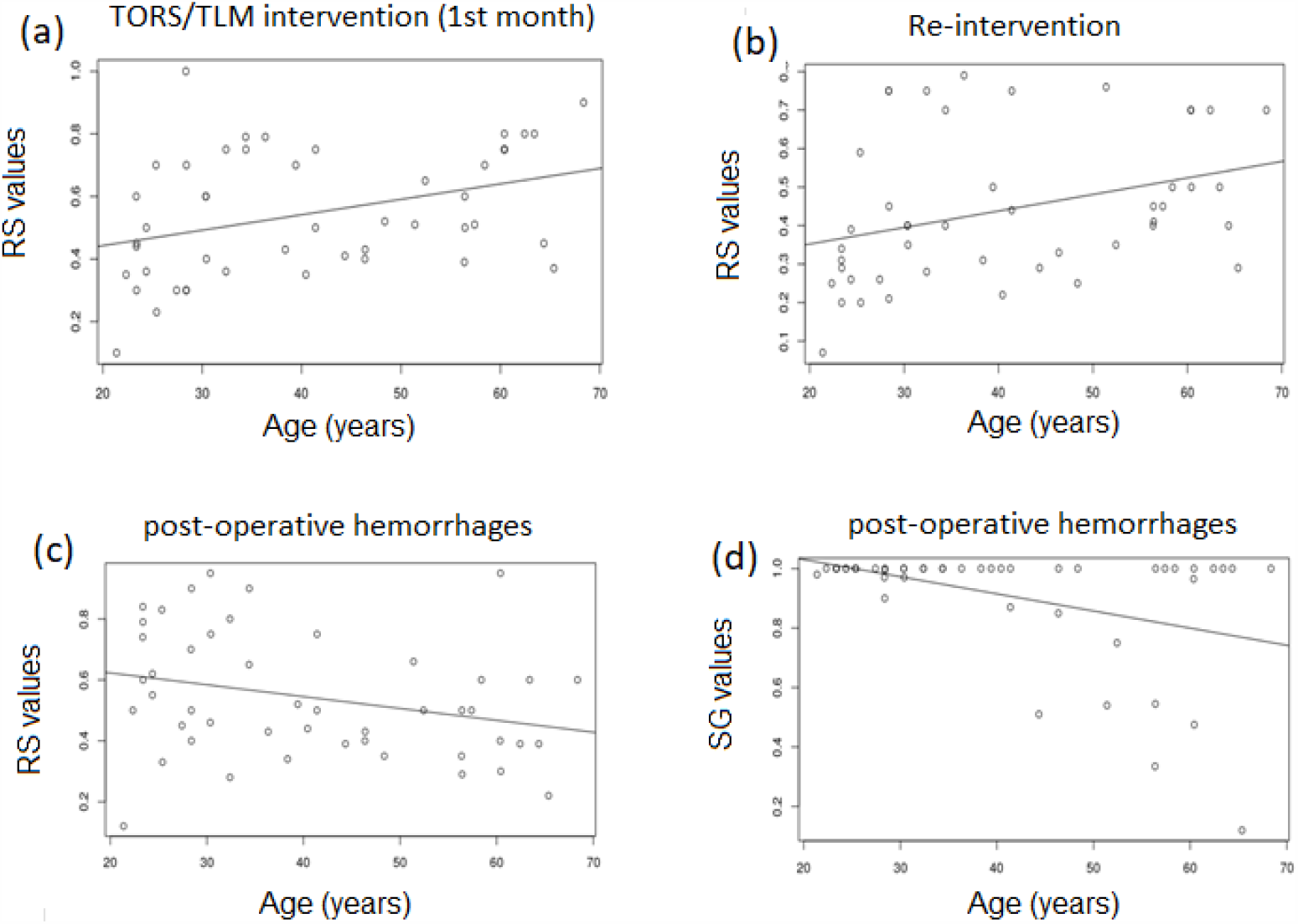
Correlation plots of RS values (a,b,c) and SG utilities (d) with age for scenarios 1 (a),2(b) and 11 (c,d).

In order to assess the need for tailoring decision models according to some population characteristics, we investigated the correlation of the elicited values with the interviewees’ profile variables (gender, age, profession, marital status, education level).

No significant difference was found between males and females (Wilcoxon rank sum test p-value: 0.1824)

About age, no significant correlation was found with UCs elicited with SG (unless a slight negative correlation for the state 11-post-operative hemorrhages, p-value = 0.08338, rho −0.255219).

Considering RS values, significant direct correlations were found with age for states 1 (p-value = 0.002363, rho 0.4330367) and 2 (p-value = 0.002985, rho 0.4282678), while significant negative correlation was found for state 11 (p-value = 0.0456, rho −0.2930534), giving support to the above finding with SG.

When considering profession, unemployed people show significantly lower UCs for 4 health states (1,2,8 and 11). While people employed in commercial activities show significantly lower UCs for 3 health states (1,8 and 12). No other significant correlations were found between profession and SG utilities. Concerning RS only for state 2 a significant positive correlation with age was found.

Regarding marital status, the only nearly significant difference was found for state 17 with lower SG UC coefficients elicited from married people. No difference were found when considering RS values.

No significant differences were found when comparing elicited values in people with different education level.

### 4.3 Correlation of elicited values with literature results

Table 5 summarizes the comparison with UCs elicited in the present study and the study by de Almeida [8] on a Canadian population that, as mentioned in section 3, is the only study reporting UCs comparable with our ones. The most notable differences are found in the least-preferable health states (e.g. distant recurrence and palliative care) where the UCs elicited in our study are lower than the ones in the de Almeida study. In particular, the 95% CIs of these last 2 health states do not overlap with the ones observed in our study, indicating a significant statistical difference (p<0.05).

**Table 5.**
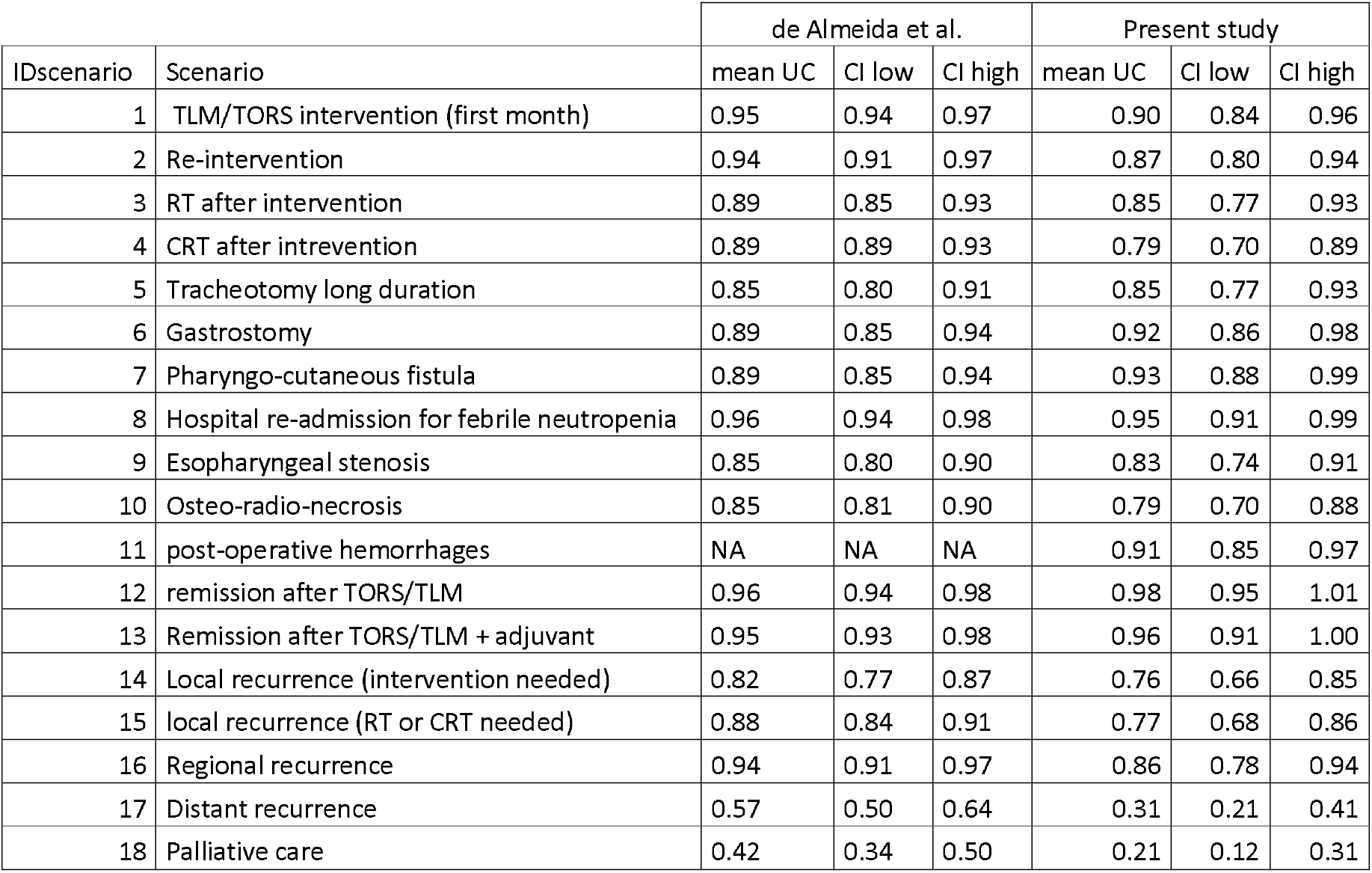
comparison of the UCs elicited with SG method in the present study and the study from de Almeida et al [8]. Mean and 95% CI are reported

| IDscenario | Scenario | de Almeida et al. |  |  | Present study |  |  |
| --- | --- | --- | --- | --- | --- | --- | --- |
|  |  | mean UC | CI low | CI high | mean UC | CI low | CI high |
| 1 | TLM/TORS intervention (first month) | 0.95 | 0.94 | 0.97 | 0.90 | 0.84 | 0.96 |
| 2 | Re-intervention | 0.94 | 0.91 | 0.97 | 0.87 | 0.80 | 0.94 |
| 3 | RT after intervention | 0.89 | 0.85 | 0.93 | 0.85 | 0.77 | 0.93 |
| 4 | CRT after intervention | 0.89 | 0.89 | 0.93 | 0.79 | 0.70 | 0.89 |
| 5 | Tracheotomy long duration | 0.85 | 0.80 | 0.91 | 0.85 | 0.77 | 0.93 |
| 6 | Gastrostomy | 0.89 | 0.85 | 0.94 | 0.92 | 0.86 | 0.98 |
| 7 | Pharyngo-cutaneous fistula | 0.89 | 0.85 | 0.94 | 0.93 | 0.88 | 0.99 |
| 8 | Hospital re-admission for febrile neutropenia | 0.96 | 0.94 | 0.98 | 0.95 | 0.91 | 0.99 |
| 9 | Esopharyngeal stenosis | 0.85 | 0.80 | 0.90 | 0.83 | 0.74 | 0.91 |
| 10 | Osteo-radio-necrosis | 0.85 | 0.81 | 0.90 | 0.79 | 0.70 | 0.88 |
| 11 | post-operative hemorrhages | NA | NA | NA | 0.91 | 0.85 | 0.97 |
| 12 | remission after TORS/TLM | 0.96 | 0.94 | 0.98 | 0.98 | 0.95 | 1.01 |
| 13 | Remission after TORS/TLM + adjuvant | 0.95 | 0.93 | 0.98 | 0.96 | 0.91 | 1.00 |
| 14 | Local recurrence (intervention needed) | 0.82 | 0.77 | 0.87 | 0.76 | 0.66 | 0.85 |
| 15 | local recurrence (RT or CRT needed) | 0.88 | 0.84 | 0.91 | 0.77 | 0.68 | 0.86 |
| 16 | Regional recurrence | 0.94 | 0.91 | 0.97 | 0.86 | 0.78 | 0.94 |
| 17 | Distant recurrence | 0.57 | 0.50 | 0.64 | 0.31 | 0.21 | 0.41 |
| 18 | Palliative care | 0.42 | 0.34 | 0.50 | 0.21 | 0.12 | 0.31 |

### 4.4 Limitations

Our study has some limitations. Confidence intervals for elicited UCs are wider than the ones reported in de Almeida [8]. This might suggest incrementing the number of interviewees, which would also allow performing sub-group analyses that may reveal more uniform opinions. Finally, our interviewees had, in large part (36/47), a high level of instruction. Thus, a not negligible selection bias has to be taken into account.

## 5. Conclusion

To the best of our knowledge this is the first study collecting UCs after head&neck surgery in Switzerland. Moreover our study collected, for the first time, UCs on health states associated to both TORS and TLM.

The comparison performed with the study from de Almeida and colleagues [8] shows similar UCs between the Canadian and Swiss population, suggesting that UCs elicited in the two studies may be valid for a broader population than the Swiss and Canadian only. At the same time, it highlighted a significant difference in the most unfavorable health states, suggesting a different perception of those states in the two countries.

The elicited values will be used first of all in the decision model for the cost/utility analysis of TORS vs TLM from the perspective of the Swiss Health National System. Beyond the specific application, the elicited values represent useful data for further economic evaluations that will consider the same health states, some of which are general enough to be applied to different cancer-related scenarios.

## Data Availability

Results data (utility coefficients) are available upon request.
The computerized elicitation software used, and the analyses code are available upon request.

## Declarations

### Availability of data and material (data transparency)

Results data (utility coefficients) are available upon request.

### Code availability (software application or custom code)

The computerized elicitation software used, and the analyses code are available upon request.

### Authors’ contributions

(optional: please review the submission guidelines from the journal whether statements are mandatory)

## Ethics approval

Since no personal data related to health is collected, and the population of participants is not recruited on the basis of a diagnosis, the research project does not fall within the scope of application of the LRH (loi fédérale relative à la recherche sur l’être humain) and does not require authorization from the CER-VD (commision ethique de la recherche de l’être humain Suisse du Canton de Vaud).

*2*.*04*.*2019 - Secrétariat scientifique, Commission cantonale (VD) d’éthique de la recherche sur l’être humain, Av. de Chailly 23, 1012 Lausanne,+4121 3161836*

Utilities represent useful data for further economic evaluations that consider the health states described in Table 2, some of which are general enough to be applied in different cancer-related scenarios.

